# The Preventive Treatment of Migraine with Low-Dose naltrexone and acetaminophen Combination: Findings of a Small, Randomized, Double-Blind, and Placebo-Controlled Clinical Trial with an Open-Label Extension for None-Responders

**DOI:** 10.1101/2021.03.23.21254186

**Authors:** Annette C. Toledano

## Abstract

We tested a low-dose naltrexone and acetaminophen combination for episodic migraine prevention. We randomly assigned patients to naltrexone and acetaminophen combination (n=6) or placebo (n=6) for a 12-week double-blind treatment. Non-responders continued into open-label treatment with naltrexone and acetaminophen combination (n=5) for additional 12 weeks. Patients were adults who experienced 5 to 17 (average 9.7) migraine days at baseline. The primary endpoint was the mean change in the monthly migraine days during the last 4 weeks of the double-blind treatment period. The key secondary endpoint was the mean change in the monthly migraine days from the 4-week double-blind follow-up (2nd baseline) to the last 4 weeks of the open-label treatment period.

The magnitude of the treatment effect for the naltrexone and acetaminophen combination observed in the double-blind period was 2.2 fewer monthly migraine days than placebo (p=0.43). Four out of 6 (66.7%) naltrexone and acetaminophen-treated patients experienced 75% reduction in migraine days compared to 1 out of 6 (16.7%) placebo-treated patients (p=0.09).

In the open-label phase, treatment with the naltrexone and acetaminophen combination (n=5) led to 8.2 fewer mean monthly migraine days (from 11.8 to 3.6), representing 69.5% improvement (p=0.03), and 100% of the patients experienced a 50% reduction in monthly migraine days. Adverse events were mild to moderate and transient, included dry mouth, fatigue, sedation, nausea, and feeling jittery.

We postulate that naltrexone’s toll-like receptor (TLR4) antagonism properties prevent pro-inflammatory cytokines’ production in the trigeminal ganglion averting “overactive nerves” (layman’s term) and migraine. Although this trial used low-dose naltrexone (defined as 1 – 5 mg/day), in future phase 3 studies we will test a range of naltrexone and acetaminophen combination doses.

## INTRODUCTION

The estimated global prevalence of migraine is 14.7% (that’s around 1 in 7 people).^1^ In the global burden of disease study, updated in 2013, migraine was the sixth highest cause worldwide of years lost due to disability (YLD).^2^ In the United States, approximately 38 million Americans are afflicted with migraine, and available treatments do not adequately meet the needs of many. Hence, there is a need for new treatments for preventing migraine. Oral naltrexone and acetaminophen combination, if proven effective, may provide greater efficacy/tolerability ratio than existing migraine prevention treatments.

Naltrexone,^3^ an opioid antagonist approved for addiction treatment, is also an analgesic due to toll-like receptor 4 (TLR4) antagonism properties. Inhibiting the TLR4 with naltrexone in nerve cells of the dorsal root ganglia (DRG) and trigeminal ganglion led to reduction in pro-inflammatory cytokines’ production (calcitonin gene-related peptide [CGRP], TNF-α, and IL-1β)^5^ and reversal of neuropathic pain and migraine in animal studies.^4–10^ Naltrexone can prevent a “localized cytokine storm” (our term) in neurons averting pain. We postulate that naltrexone’s toll-like receptor (TLR4) antagonism properties prevent pro-inflammatory cytokines’ production in the trigeminal ganglion averting “overactive nerves” (layman’s term) and migraine.

The Interagency Pain Research Coordinating Committee (IPRCC) voted a study that used naloxone^5^ (a opioid/TLR4 antagonist similar to naltrexone) as one of 2009-2013 pain research advances that represent significant progress in the field. “This research supports TLR4 as a potential therapeutic target for treating chronic pain in patients, and, as the establishment of a completely new class of pain-relieving medication, would be a remarkable advance in pain treatment.”^11^

TLR4 is an innate immune system receptor that usually detects invasion of foreign agents such as viruses and initiates a cascade leading to cytokines’ production to eliminate them. However, the TLR4 can also be triggered by endogenous damage molecules originating from injured tissues (e.g., a herniated intervertebral disc) and lead to cytokines’ production and pain.^12^ The inborn, innate immune system is not to be confused with the learned, adaptive immune system.

Naltrexone’s analgesic properties are due to its inhibition of the production of pro-inflammatory cytokines, e.g., interleukin (IL)-6, tumor necrosis factor (TNF)-α, interferon-β, calcitonin gene-related peptide (CGRP), nitric oxide (NO), and reactive oxygen species (ROS) in nerve cells, ^5,8,13,14^ preventing a “localized cytokine storm” (our term) and pain generation. Naltrexone exerts its action at the beginning of the cascade leading to the production of many pro-inflammatory cytokines blocking their creation. Naltrexone can prevent the production of multiple cytokines eliminating the need to neutralize them after they were created.

We hypothesize that the pathophysiological event underlying migraine is the excessive production of pro-inflammatory cytokines in the trigeminal ganglion creating a neuro-inflammatory response resulting in a “localized cytokine storm.” Similarly, in the dorsal root ganglions, a “localized cytokine storm” results in neuropathic back pain. We postulate that a localized “cytokine storm” is the underlying event leading to “overactive nerves” (layman’s term) and neuropathic pain.

Pro-inflammatory cytokines exaggerate neuronal excitability, contributing to neuropathic pain and migraine. Activation of TLR4 has been implicated in the pathogenesis of migraine^9,15^ and (+)-Naltrexone blocked the development of facial allodynia in modeled migraine in rats.^9^

Dr. Bernard Bihari invented Low-Dose Naltrexone (LDN) (a daily dose of 1 to 5 mg) in the mid-1980s for “normalizing the immune system function.”^16,17^ However, scientists discovered the innate immune system and TLRs in humans in the 1990s (a Nobel Prize was awarded in 2011).^18^ The prevailing theory for LDN’s mechanism of action was that it increases endorphin production, systemically upregulating endogenous opioid signaling by a transient opioid-receptor blockade.^19^ later research attributed the analgesic properties of naltrexone to TLR4 antagonism leading to pro-inflammatory cytokines’ production inhibition. Currently, LDN is widely accepted as an alternative medicine modality and is used by its proponents to treat various medical conditions. It is almost sold as an everyday supplement by certain pharmacies.^19,20^

Grassroots interest in off-label LDN sprang clinical trials for fibromyalgia, multiple sclerosis, Crohn’s disease, and complex regional pain syndrome (CRPS).^21–25^ Although LDN is reportedly used as an off-label treatment for various medical conditions, there are no confirmatory studies for these off-label uses. Addiction specialists are the primary prescribers of naltrexone.

Acetaminophen potentially enhances the combination in several ways. Acetaminophen created synergy for analgesia in combination drugs such as Vicodin (hydrocodone/ acetaminophen).^26^ Similarly, combining naltrexone with acetaminophen could attain synergy for analgesia. Acetaminophen 1000 mg was already established as an effective acute migraine medication and can therefore enhance the combination’s treatment effect on migraine.^27,28^ Acetaminophen has the public’s trust as the world’s most consumed drug. Acetaminophen was found to be also an emotional pain reducer. Acetaminophen significantly reduced hurt feelings in human studies.^29,30^ Acetaminophen’s emotional pain reducing properties could potentially be augmented by naltrexone^31^ and enhance the combination’s effect on the overall sense of well-being.

Naltrexone’s cytokine-production inhibition properties in the trigeminal and dorsal root ganglia leading to reversal of pain could reduce pain in COVID-19 and COVID-19 vaccination. Three experimental TLR4 antagonists FP7, Eritoran, and retrocyclin 101, were significantly better than placebo in treating lethal influenza.^32–34^ TLR4 signaling is a key disease pathway controlling the severity of acute lung injury.^35^ Naltrexone, a readily available TLR4 antagonist, needs to be evaluated for the treatment of pain symptoms in COVID-19.

Interestingly, the innate immune system’s pathway that mounts an initial response to viral infections is the same one that leads to neuropathic pain when triggered intrinsically by damage-molecules.

Naltrexone and acetaminophen combines naltrexone’s unutilized analgesic properties with the established and well-trusted analgesic properties of acetaminophen.

## METHODS

### Study Population

Patients were recruited to our single site in Miami, Florida, through billboards and social media advertising. The study was conducted between August 25, 2017, and July 26, 2018. ANODYNE-3, (the study’s name) enrolled patients 18 to 75 years of age with a history of migraine with or without aura for at least one year that was consistent with the diagnosis criteria of the International Classification of Headache Disorders (ICHD)-3rd edition (beta version)^36^ and have experienced 5 to 17 migraine days in the 4-week baseline. The study population included 11 patients with episodic migraine and one with chronic migraine. Patients needed to have at least 5 migraine days during the baseline. Patients using opioid medications or who had a history within the previous 3 years of drug abuse were excluded.

Our goal was to enroll 48 patients, but we were unable to meet that goal due to hurricane Irma’s (September 2017) impact on our community.

### Trial Oversight

The sponsor/investigator – Annette Toledano, M.D., was responsible for all the trial elements, including design, execution, data collection, analysis, and interpretation.

The trial’s protocol and informed consent were approved by the Schulman Associates IRB (now Advarra). All patients provided written informed consent before starting the study procedures. The informed consent informed the patients of the conflict of interest of the sponsor/investigator. The study patients were compensated for participation.

This study was conducted under an Investigational New Drug (IND) application with the United States Food and Drug Administration (FDA).

### Trial Design

The ANODYNE-3 trial was a phase 2, single-site, randomized, double-blind, and placebo-controlled proof-of-concept study. We evaluated the patients in seven site visits during the double-blind phase and four visits during the open-label phase. The visits included: baseline (week −4), randomization (week 0), and double-blind treatment (weeks 1, 4, 8, and 12), follow-up (week 16), open-label treatment (weeks 20, 24, and 28), and open-label follow-up (week 32) (Figure 1).

**Figure 1:**
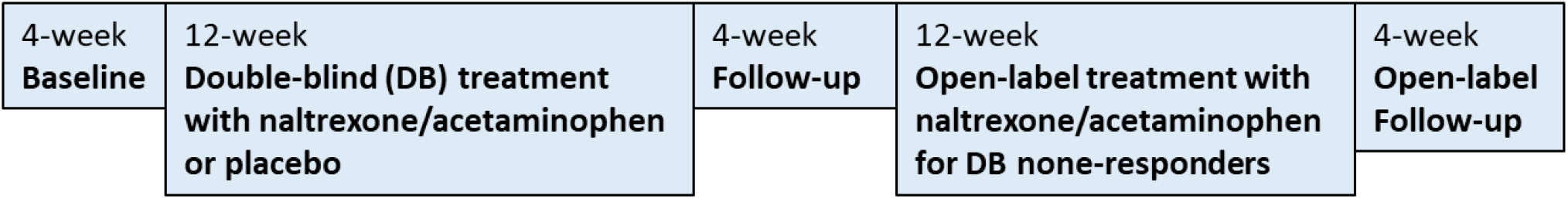
Study design flow-chart

We randomly assigned patients in 1:1 ratio to naltrexone 2.25 mg/acetaminophen 325 mg (n=6) or placebo (n=6) BID for a 12-week double-blind treatment period. Non-responders (defined as ≤50% reduction in monthly migraine days in the last 4 weeks of treatment) continued into open-label naltrexone and acetaminophen treatment (n=5) for additional 12 weeks. The open-label phase intended to gain insight into a “real world” response to the drug.

Patients recorded in a daily paper diary the duration and headache severity. They also recorded photophobia, phonophobia, nausea, vomiting, and any acute migraine medication use.

The study pharmacist prepared the study medication from marketed tablets placed in two single-ingredient capsules backfilled with microcrystalline cellulose. The study pharmacist devised the randomization schedule using a block size of 12. The study medication kits were sequentially numbered, and the investigator assigned the kits to patients consecutively. Patients took the study medication at approximately 8 am and 8 pm every day. At each site visit, the patients received enough medication to cover until the next visit, plus three extra days in case of a delayed appointment. Patients returned bottles and extra capsules at each site visit for drug accountability.

### Efficacy Assessments and Endpoint

The primary efficacy endpoint was the mean change in monthly migraine days from baseline to the last 4 weeks of the double-blind treatment period. The key secondary efficacy endpoint was the mean change in monthly migraine days from the double-blind follow-up (2nd baseline) to the last 4 weeks of the open-label treatment period.

Additional secondary endpoints were the proportion of patients achieving at least 50%, 75%, and 100% reduction in monthly migraine days, acute migraine medication treatment days, migraine headache-hours, at least moderate migraines days, and severe migraine days from baseline to the last 4 weeks of treatment. Other endpoints were Headache Impact Test (HIT-6), Pittsburgh Insomnia Rating Scale-20 (PIRS-20), and Patient Global Impression of Change (PGIC). We assessed treatment-emergent adverse events and serious adverse events. We performed a physical examination, obtained vital signs (systolic and diastolic blood pressure, pulse, and weight), and recorded concomitant medications during each site visit. The patients had a 12-lead electrocardiogram at the screening visit and monthly clinical laboratory tests (serum chemistry and hematology).

### Statistical Analysis

The efficacy and safety analyses included all randomized patients who received at least one dose of the study medication, recorded a baseline migraine severity rating, and reported at least one post-dose assessment. All statistical tests were 2-sided hypothesis tests performed at the 5% significance level. Statistical analyses were conducted using MedCalc statistical software.^37^

## RESULTS

### Patients

We assessed 94 patients for eligibility; 12 patients met the inclusion criteria and were randomized. Six patients received naltrexone 2.25 mg/acetaminophen 325 mg and 6 received placebo. All the 12 randomized patients completed the double-blind treatment period, and 5 of 6 non-responders completed the open-label treatment period (Figure 2).

**Figure 2:**
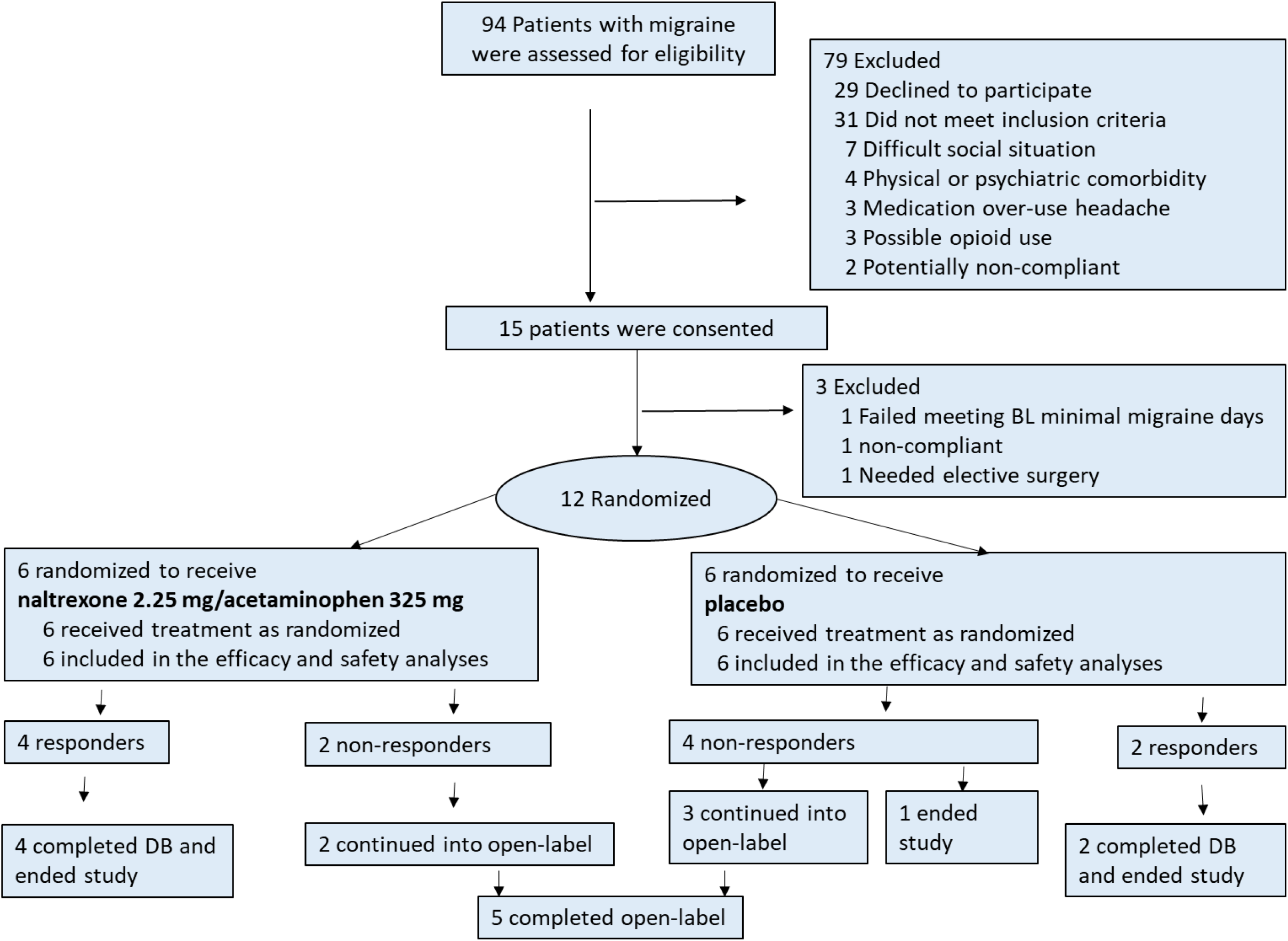
Patients’ distribution

The baseline characteristics of the patients were similar in the two groups (Table 1). The mean age was 40.3 years, and the trial population was predominantly female (83.3%) and Caucasian (91.7%). Migraine histories were similar in terms of migraine duration, the number of migraine days per month in the previous 3 months, and the monthly migraine days with acute medication use over the preceding 3 months. Acute migraine medication classes, specifically triptans – were identical in both groups. Migraine characteristics were similar among the treatment groups during the baseline period. The mean number of headache days in the 4-week baseline was 9.7 for both groups and the days of acute migraine medication use were 8.2 and 7.7 for naltrexone/ acetaminophen and placebo, respectively. Headache impact test (HIT) and Pittsburgh Insomnia Rating Scale-20 (PIRS-20) scores were comparable at the screening visit.

**Table 1:** Baseline characteristics

|  | Total<br>(n=12) | Naltrexone<br>2.25 mg/<br>acetaminophe<br>n 325 mg<br>(n=6) | Placebo<br>(n=6) |
| --- | --- | --- | --- |
| Characteristics |  |  |  |
| Age - years | 40.3±12.0 | 41.5±14.3 | 39.0±10.4 |
| Female gender - no. (%) | 10 (83.3) | 4 (66.7) | 6 (100) |
| Race or ethnic group - no. (%)† |  |  |  |
| Caucasian, no. (%) | 11 (91.7) | 6 (100.0) | 5 (83.3) |
| African American, no. (%) | 1 (8.3) | 0 (0.0) | 1 (16.7) |
| Hispanic ethnic group, no. (%) | 8 (66.7) | 4 (66.7) | 4 (66.7) |
| Body mass index‡ | 26.8±7.0 | 26.9±6.0 | 26.7±8.4 |
| Migraine History |  |  |  |
| Duration of migraine illness - year | 17.0±13.1 | 20.3±12.4 | 13.7±13.8 |
| Migraine days/mth in past 3 months – no. | 8.9±1.9 | 8.8±2.3 | 9.0±1.5 |
| Migraine days/month with acute medication use in past 3 months, no. | 8.5±2.2 | 8.2±2.6 | 8.8±1.9 |
| Previous use of preventive medications - no. % | 3 (25.0) | 1 (16.7) | 2 (33.3) |
| Acute migraine medication classes |  |  |  |
| Combination analgesics - no. (%) | 5 (41.7) | 2 (33.3) | 3 (50.0) |
| Non-steroidal anti-inflammatory drugs - no. (%) | 2 (16.7) | 2 (33.3) | 0 (0.0) |
| Acetaminophen - no. (%) | 1 (8.3) | 0 (0.0) | 1 (16.7) |
| Triptans - no. (%) | 4 (33.3) | 2 (33.3) | 2 (33.3) |
| Migraine characteristics during baseline |  |  |  |
| Migraine days per month§ | 9.7±3.3 | 9.7±4.2 | 9.7±2.9 |
| Acute migraine medication days/month | 8.2±2.7 | 7.7±2.1 | 8.7±3.5 |
| Monthly migraine hours, avg./patient | 62.6±41.2 | 70.8±47.6 | 54.4±36.2 |
| Headache Impact Test (HIT-6) score at screening¶ | 70.2±5.2 | 70.0±6.3 | 70.2±4.9 |
| Pittsburgh Insomnia Rating Scale-20 (PIRS-20) score at screening** | 32.0±13.0 | 35.7±13.9 | 27.2±13.3 |
| *Plus-minus values are means ±SD<br>†Race and ethnic group were reported by the patient.<br>‡Calculated as weight in kilograms divided by height in meters squared.<br>§ A migraine day was defined as a calendar day in which a patient reported a headache of ≥ 30-minute duration and met criteria for migraine with or without aura.<br>¶ Headache Impact Test (HIT-6). Scores range from 36 to 78, with a score of over 50 indicating disability. **Pittsburgh Insomnia Rating Scale-20 (PIRS-20). PIRS-20 scores range from 0 to 60, with scores above 20 suggesting insomnia. |  |  |  |

### Efficacy Results

Comparing the baseline to the last 4-weeks of the double-blind period, treatment with naltrexone and acetaminophen (n=6) led to 5.7 fewer monthly migraine days, whereas treatment with placebo led to 3.5 fewer monthly migraine days, both groups improving from a baseline rate of 9.7 monthly migraine days. The magnitude of the treatment effect observed in the double-blind period was 2.2 fewer monthly migraine days (p=0.43) (Table 2 and Figure 3). In the double-blind period, 66.7% of the naltrexone and acetaminophen patients experienced a 50% reduction in migraine headache days from baseline to the last 4 weeks of the double-blind period compared to 33.3% in placebo patients (p=0.27). 66.7% of naltrexone and acetaminophen patients experienced 75% reduction in migraine days compared to 16.7% of placebo patients, (p=0.09), and 50.0% of the naltrexone and acetaminophen-treated patients experienced 100% reduction in monthly migraine days compared to 16.7% of placebo patients, (p=0.27) (Table 2).

**Table 2:**
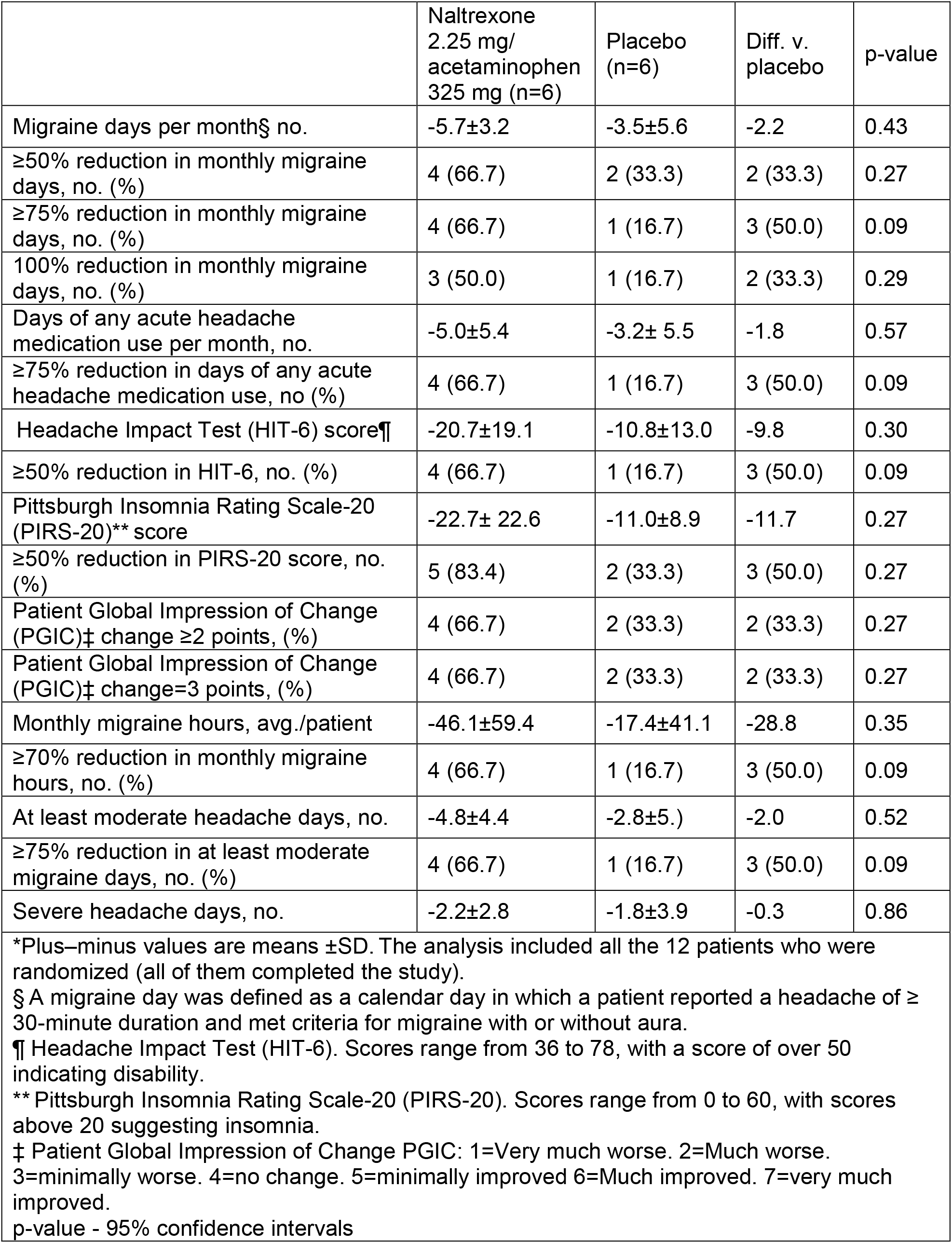
Endpoints from baseline to month-3 of the double-blind treatment period*

|  | Naltrexone<br>2.25 mg/<br>acetaminophen<br>325 mg (n=6) | Placebo<br>(n=6) | Diff. v.<br>placebo | p-value |
| --- | --- | --- | --- | --- |
| Migraine days per month§ no. | -5.7±3.2 | -3.5±5.6 | -2.2 | 0.43 |
| ≥50% reduction in monthly migraine days, no. (%) | 4 (66.7) | 2 (33.3) | 2 (33.3) | 0.27 |
| ≥75% reduction in monthly migraine days, no. (%) | 4 (66.7) | 1 (16.7) | 3 (50.0) | 0.09 |
| 100% reduction in monthly migraine days, no. (%) | 3 (50.0) | 1 (16.7) | 2 (33.3) | 0.29 |
| Days of any acute headache medication use per month, no. | -5.0±5.4 | -3.2± 5.5 | -1.8 | 0.57 |
| ≥75% reduction in days of any acute headache medication use, no (%) | 4 (66.7) | 1 (16.7) | 3 (50.0) | 0.09 |
| Headache Impact Test (HIT-6) score¶ | -20.7±19.1 | -10.8±13.0 | -9.8 | 0.30 |
| ≥50% reduction in HIT-6, no. (%) | 4 (66.7) | 1 (16.7) | 3 (50.0) | 0.09 |
| Pittsburgh Insomnia Rating Scale-20 (PIRS-20)** score | -22.7± 22.6 | -11.0±8.9 | -11.7 | 0.27 |
| ≥50% reduction in PIRS-20 score, no. (%) | 5 (83.4) | 2 (33.3) | 3 (50.0) | 0.27 |
| Patient Global Impression of Change (PGIC)‡ change ≥2 points, (%) | 4 (66.7) | 2 (33.3) | 2 (33.3) | 0.27 |
| Patient Global Impression of Change (PGIC)‡ change=3 points, (%) | 4 (66.7) | 2 (33.3) | 2 (33.3) | 0.27 |
| Monthly migraine hours, avg./patient | -46.1±59.4 | -17.4±41.1 | -28.8 | 0.35 |
| ≥70% reduction in monthly migraine hours, no. (%) | 4 (66.7) | 1 (16.7) | 3 (50.0) | 0.09 |
| At least moderate headache days, no. | -4.8±4.4 | -2.8±5.) | -2.0 | 0.52 |
| ≥75% reduction in at least moderate migraine days, no. (%) | 4 (66.7) | 1 (16.7) | 3 (50.0) | 0.09 |
| Severe headache days, no. | -2.2±2.8 | -1.8±3.9 | -0.3 | 0.86 |
| *Plus-minus values are means ±SD. The analysis included all the 12 patients who were randomized (all of them completed the study).<br>§ A migraine day was defined as a calendar day in which a patient reported a headache of ≥ 30-minute duration and met criteria for migraine with or without aura.<br>¶ Headache Impact Test (HIT-6). Scores range from 36 to 78, with a score of over 50 indicating disability.<br>** Pittsburgh Insomnia Rating Scale-20 (PIRS-20). Scores range from 0 to 60, with scores above 20 suggesting insomnia.<br>‡ Patient Global Impression of Change PGIC: 1=Very much worse. 2=Much worse. 3=minimally worse. 4=no change. 5=minimally improved 6=Much improved. 7=very much improved.<br>p-value - 95% confidence intervals |  |  |  |  |

**Figure 3:**
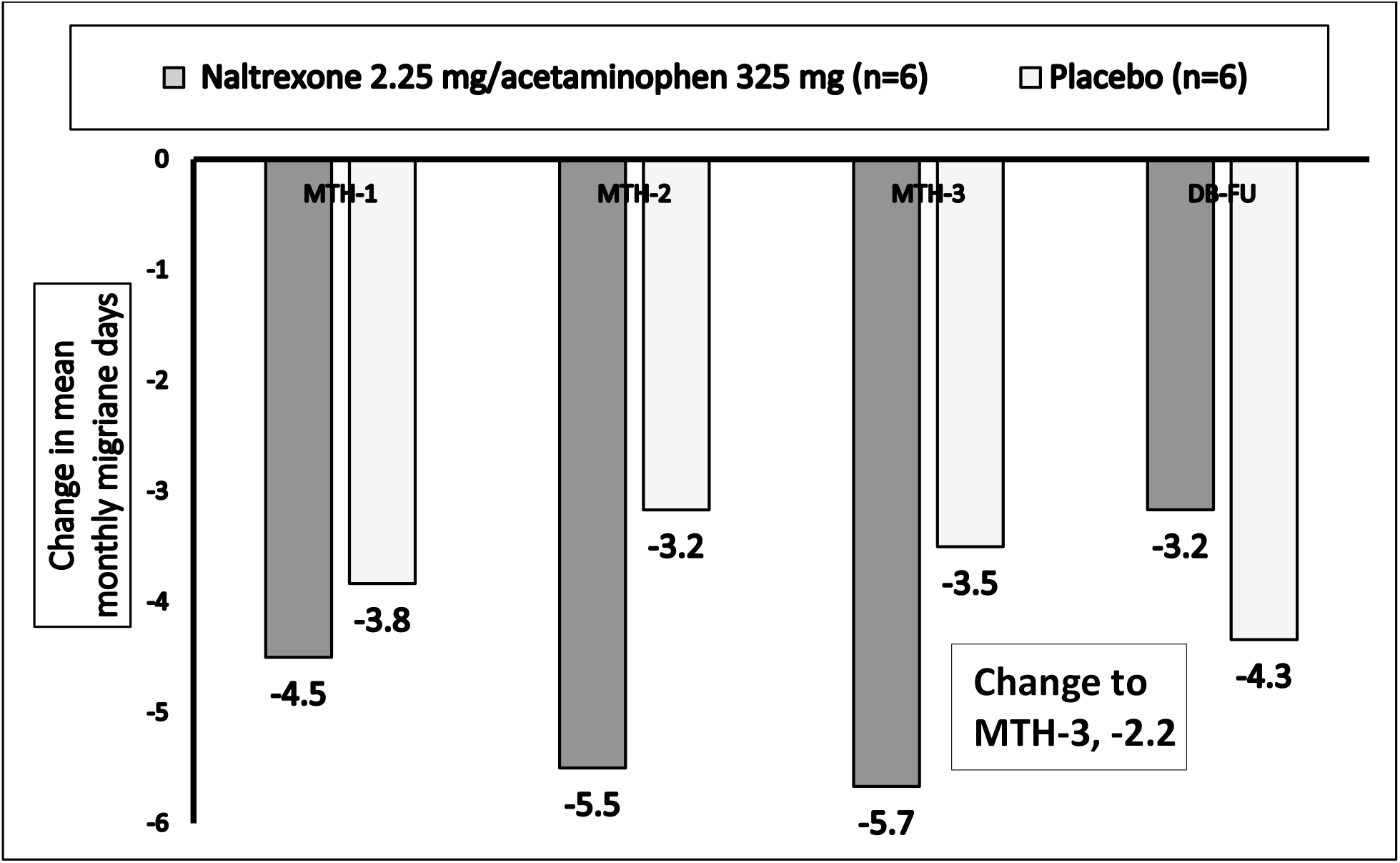
Change in mean monthly migraine days from baseline in the double-blind phase.

The magnitude of the treatment effect from baseline to the last 4 weeks of the double-blind treatment for Headache Impact Test (HIT) score was −9.8 points (p=0.29), and for Pittsburgh Insomnia Rating Scale-20, it was −11.7 (p=0.27). The magnitude of the treatment effect in monthly migraine hours in the double-blind period was a 28.8 hours reduction (p=0.35).

In the open-label period, treatment with naltrexone and acetaminophen (n=5) led to 8.20 fewer monthly migraine days (from 11.80 to 3.60), amounting to 69.5% improvement, (p=0.03) (Figure 4). Also, 100% of the patients experienced a 50% reduction in the monthly migraine days (Table 3). The treatment effect from the double-blind follow-up to the last 4 weeks of the open-label period for Headache Impact Test and Pittsburgh Insomnia Rating Scale-20 was −25.8 (p=0.0002) and −17.0 (p=0.09) points, respectively. The treatment effect in monthly migraine hours in the open-label period was −36.2 hours (p=0.09). In the open-label period, there were statistically significant reductions from baseline to the last 4 weeks of treatment in the days of use of any acute headache medications, Headache Impact Test (HIT), at least moderate migraine days, and severe headache days.

**Table 3:** Endpoints from double-blind follow-up (2^nd^ baseline) to month-3 of open-label treatment period*

|  | Double-blind follow-up (baseline) (n=5) | last 4 weeks of open-label treatment (n=5) | Difference from baseline | p-value |
| --- | --- | --- | --- | --- |
| Migraine days per month, § no. | 11.8±6.0 | 3.6±3.7 | -8.2±3.9 | 0.03 |
| ≥50% reduction in monthly migraine days, no. (%) |  | 5 (100) | 5 (100) | n/a |
| ≥75% reduction in monthly migraine days, no. (%) |  | 3 (60) | 3 (60) | n/a |
| 100% reduction in monthly migraine days, no. (%) |  | 3 (60) | 3 (60) | n/a |
| Days of use of any acute headache medications per month, no. | 9.8±4.3 | 1.6± 2.5 | -8.2±2.8 | 0.0063 |
| Headache Impact Test (HIT-6) score¶ | 68.2±5.7 | 42.4± 7.1 | -25.8±4.4 | 0.0002 |
| HIT-6 Mean % response (adjusted for score of 36 being 100% improvement) |  |  | 80% (15%) | 0.0002 |
| ** Pittsburgh Insomnia Rating Scale-20 (PIRS-20) score | 25.0±18.5 | 8.0±7.0 | -17.0±17.2 | 0.09 |
| Patient Global Impression of Change (PGIC)‡ change ≥2 points, (%) |  | 5, (100) | 5, (100) | n/a |
| Patient Global Impression of Change (PGIC)‡ change=3 points, (%) |  | 4, (80) | 4, (80) | n/a |
| Monthly migraine hours, no. | 55.3±34.5 | 19.2±24.6 | -36.2±32.4 | 0.09 |
| At least moderate migraine days, no. | 10.8±5.0 | 2.6±2.7 | -8.1±4.2 | 0.01 |
| Severe headache days, no. | 6.8±4.8 | 1.0±1.2 | -5.8±3.4 | 0.03 |
| * Plus-minus values are means ±SD. The analysis included 5 of the 6 patients who were non-responders in the double-blind period. Non-responders were defined as ≤50% reduction in monthly migraine days in the last 4 weeks of treatment.<br>§ A migraine day was defined as a calendar day in which a patient reported a headache of ≥ 30-minute duration and met criteria for migraine with or without aura.<br>¶ Headache Impact Test (HIT-6). Scores range from 36 to 78, with a score of over 50 indicating disability.<br>** Pittsburgh Insomnia Rating Scale-20 (PIRS-20). Scores range from 0 to 60, with scores above 20 suggesting insomnia.<br>‡ Patient Global Impression of Change (PGIC): 1=Very much worse. 2=Much worse. 3=minimally worse. 4=no change. 5=minimally improved 6=Much improved. 7=very much improved.<br>p-value - 95% confidence intervals |  |  |  |  |

**Figure 4:**
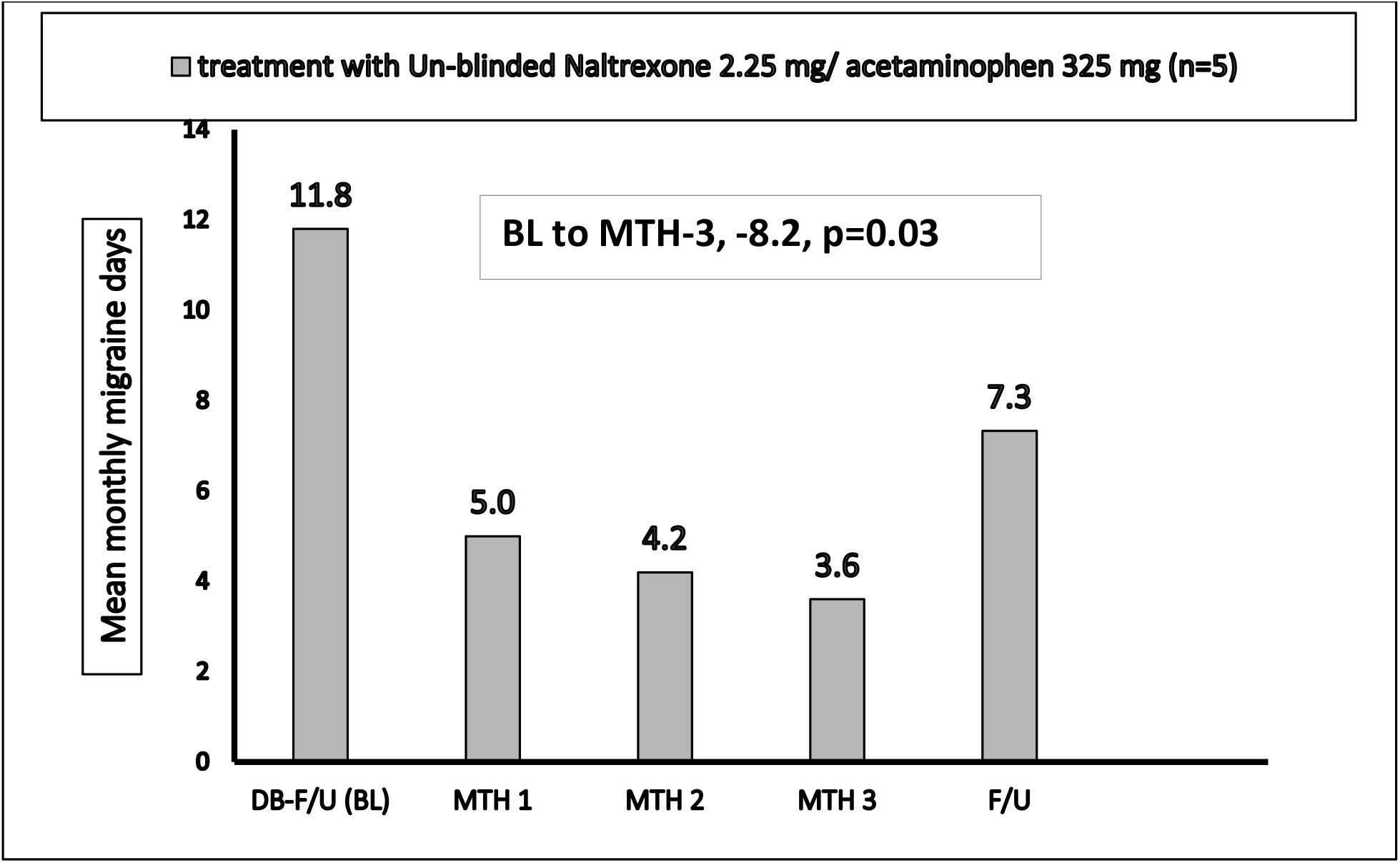
Mean monthly migraine days from baseline in the open-label phase.

The improvements in Headache Impact Test (HIT) and Pittsburgh Insomnia Rating Scale-20 (PIRS-20) indicate enhancement of quality of life measures.

Acetaminophen-overuse head could be expected in patients having more than 15 headaches per month who consume acetaminophen more than 15 days per month for more than 3 months.^36^

Since the study administered acetaminophen 650 mg daily, we were concerned that acetaminophen may induce medication overuse headaches. The study provided data on 11 three-month treatment courses with naltrexone and acetaminophen for nine unique patients and two patients who received two consecutive courses separated by 4 weeks. During each of these courses, migraine days were lower than the baseline (Figure 5 and Figure 6). This study provided preliminary evidence that a daily dose of naltrexone 4.5 mg/acetaminophen 650 mg did not induce medication overuse headache after three months of treatment.

**Figure 5:**
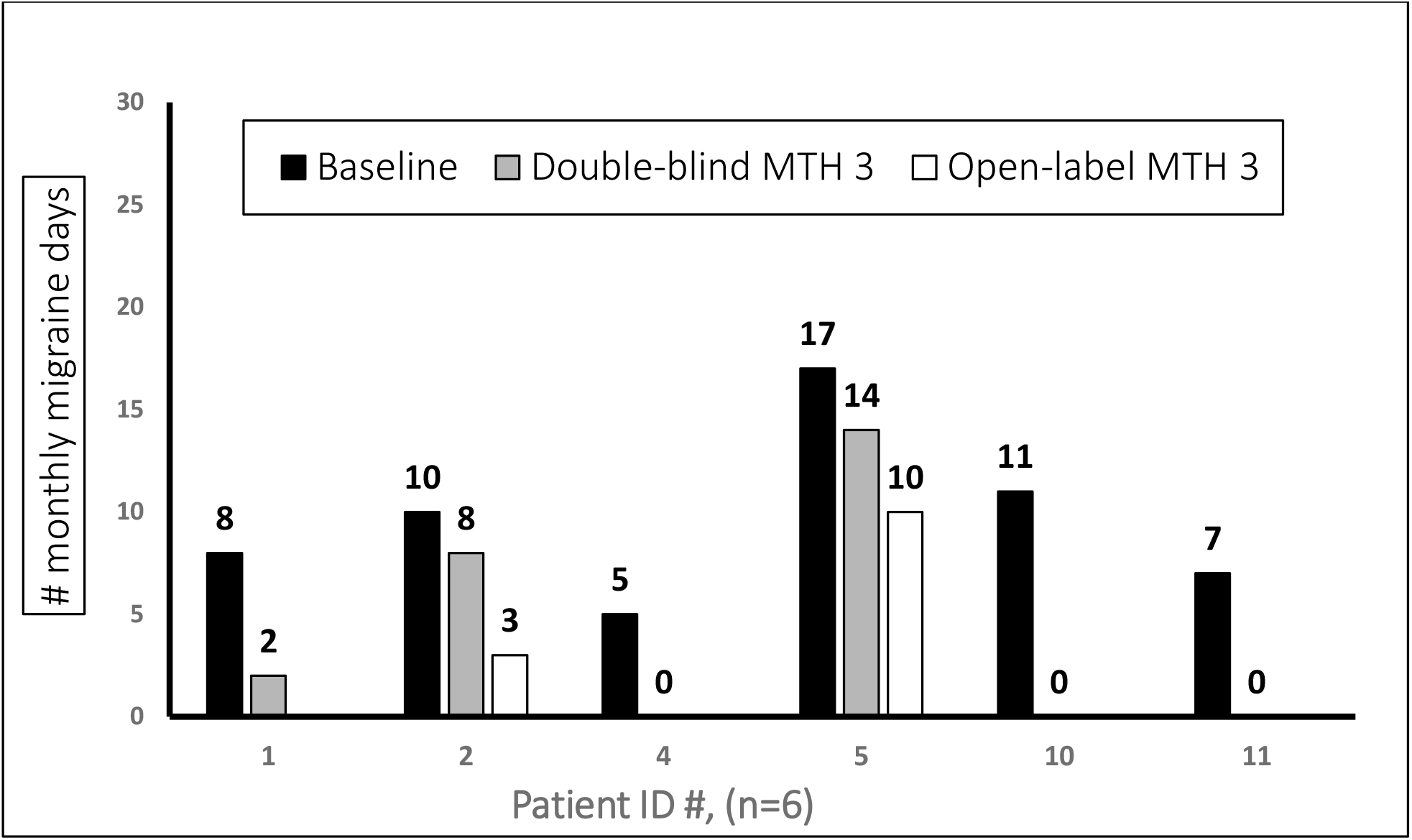
Monthly migraine days for individual naltrexone and acetaminophen-treated patients in the double-blind phase.

**Figure 6:**
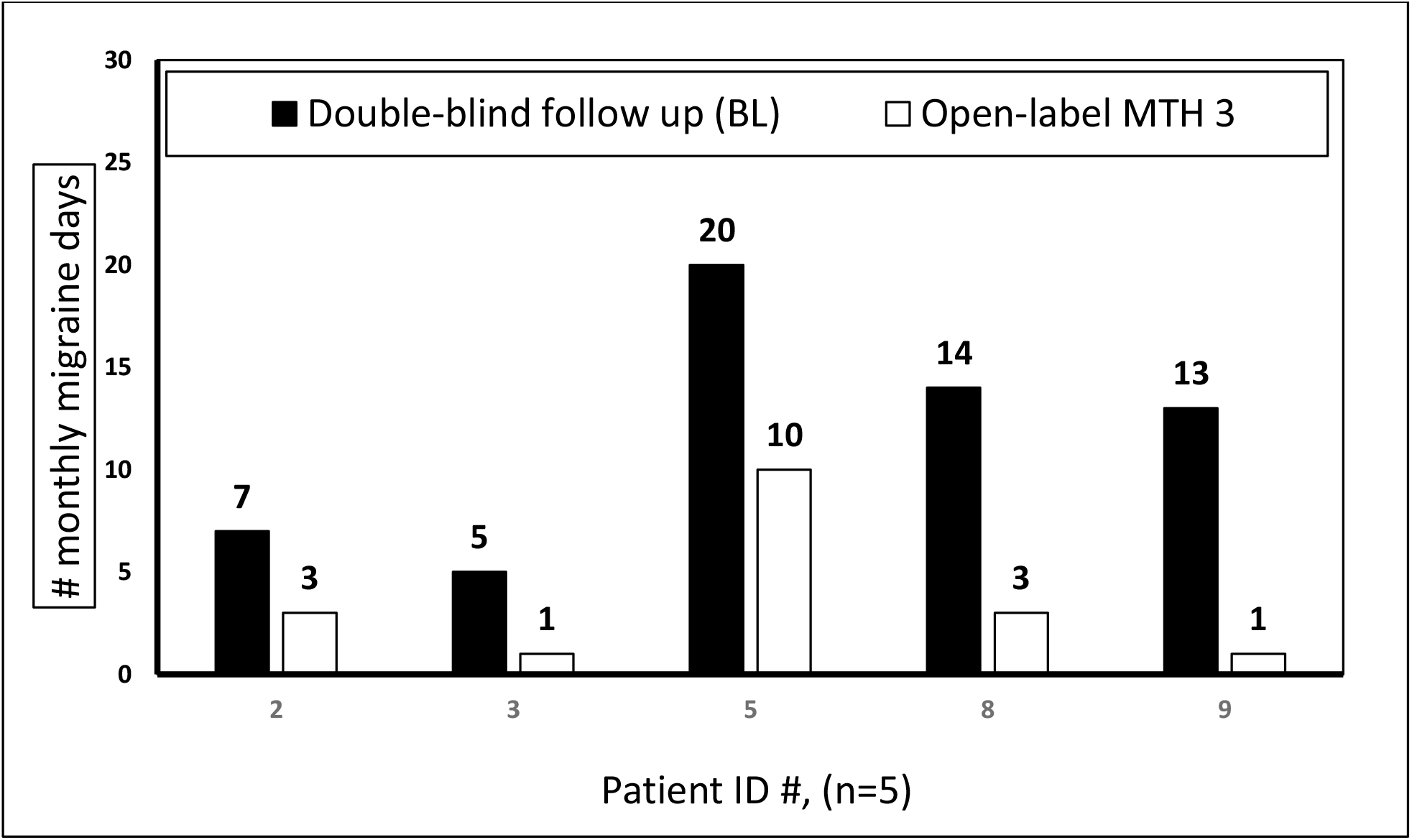
Monthly migraine days in individual naltrexone and acetaminophen-treated patients in the open-label phase. BL=Baseline.

### Safety

Treatment-emergent adverse events were reported by 66.7% (4 of 6) of patients in the naltrexone 2.25 mg/acetaminophen 325 mg group, 33.3% (2 of 6) in the placebo group, and 60.0% (3 of 5) in the open-label period. (Table 4).

**Table 4:** Treatment-emergent adverse events*

|  | Naltrexone 2.25 mg/<br>acetaminophen 325 mg<br>(n=6) | Placebo<br>(n=6) | Un-blinded Naltrexone<br>2.25 mg/ acetaminophen<br>325 mg (n=5) |
| --- | --- | --- | --- |
| Death — no. (%) | 0 | 0 | 0 |
| Serious adverse event —<br>no. (%) | 0 | 0 | 0 |
| Discontinuation due to<br>adverse event - no. (%) | 0 | 0 | 0 |
| Patients ≥1 Adverse event<br>during the trial — no. (%) | 4 (66.7) | 2 (33.3) | 3 (60) |
| All adverse events* |  |  |  |
| Dry Mouth — no. (%) | 1 (16.7) |  | 1 (20.0) |
| Fatigue — no. (%) | 1 (16.7) | 1 (16.7) | 1 (20.0) |
| Feeling Jittery |  |  | 1 (20.0) |
| Gastroesophageal reflux<br>— no. (%) | 1 (16.7) |  |  |
| Heavy breathing — no.<br>(%) | 1 (16.7) | 1 (16.7) |  |
| Nausea — no. (%) | 1 (16.7) |  |  |
| Palpitations, no. (%) | 1 (16.7) |  |  |
| Sedation, no. (%) | 1 (16.7) | 1 (16.7) | 1 (20.0) |
| Tinnitus, no. (%) |  |  | 1 (20.0) |
| Vomiting, no. (%) | 1 (16.7) |  |  |
| Total adverse events, no. | 8 | 3 | 5 |
| * All the reported adverse events. |  |  |  |

Dry mouth, fatigue, sedation, nausea, and feeling jittery were the most commonly reported events. All the adverse events were mild/moderate in severity and resolved within 1 to 3 days. No serious adverse events were reported. No deaths or discontinuations due to an adverse event were reported. No changes in monthly liver transaminases or bilirubin were observed.

## DISCUSSION

In the preventive treatment of migraine, naltrexone and acetaminophen demonstrated higher response rates than placebo in several key endpoints. The double-blind and open-label phase results together suggest naltrexone and acetaminophen may be useful in the preventive treatment of migraine.

There were fewer migraine days per month in the double-blind phase and higher proportions of patients experienced a 50%, 75%, and 100% reduction in monthly migraine days in the naltrexone and acetaminophen-treated group. Naltrexone and acetaminophen improved the quality of life compared with placebo, as indicated by the Headache Impact Test, Pittsburgh Insomnia Rating Scale-20, and the Patient Global Impression of Change.

The open-label treatment phase, representing “real world” response, demonstrated statistically significant reductions in migraine days per month, days of any acute headache medications use, Headache Impact Test score, at least moderate migraine days, and severe headache days.

The daily dose of naltrexone in this study was approximately 1/10th of the approved addiction treatment dose. The daily acetaminophen dose was 650 mg.

Although this trial used low-dose naltrexone (defined as 1 – 5 mg/day), in future phase 3 studies we will test a range of doses of the naltrexone and acetaminophen combinations.

Limitations of this trial included its tiny sample size and treatment periods of just 3 months. The treatment period needed to be longer than 3 months to assess the acetaminophen-overuse headache risk.

Targeting the toll-like receptor-4 represents a novel approach to treating migraine. Adding acetaminophen, the world’s most used drug, could enhance the combination’s analgesic effect and boost the public’s trust conferring an advantage over naltrexone alone. This trial provides preliminary evidence for the potential benefits of naltrexone as a migraine prevention treatment.

This study was the first to assess a naltrexone product for migraine prevention.

Adequately powered clinical trials are needed to confirm these findings. The study’s sample size was too small to achieve statistical significance for the primary endpoint.

## PATENTS

The naltrexone and acetaminophen combination received two U.S. patents for treating pain and a U.S. patent for treating emotional pain.

## Data Availability

Data not available

## Author Contribution

**Concept, design, statistical analysis, and data interpretation:** Annette Toledano.

Dr. Toledano had full access to all of the data in the study and takes responsibility for the integrity of the data and the accuracy of the data analysis.

**Study Pharmacist:** Ayman Mohamed PharmD.

## Conflict Of Interest Disclosures

Dr. Annette Toledano reports she is the founder/medical director of Allodynic Therapeutics, LLC and the inventor/patent holder of several naltrexone and acetaminophen patents. Allodynic Therapeutics is a clinical-stage, specialty Biopharmaceutical Company focused on painful conditions with high unmet needs.

### Funding/Support

This trial was fully funded by Allodynic Therapeutics, LLC.

### Disclaimer

The opinions expressed in this article are those of the author.

### Grants

None

### Trial Registration

ClinicalTrials.gov Identifier: NCT03194555.

### Institutinal Review Board (IRB)

This trial’s protocol and informed consent were approved by the Schulman Associates IRB (now Advarra).

## Acknowledgments

We acknowledge the immense contribution of the study patients, the study pharmacist, and the clinical research coordinators, whose hard work made this trial possible.

## Notes

### Clinical Trial

ClinicalTrials.gov Identifier: NCT03194555.

### Funding Statement

No funding

### Author Declarations

Schulman Associates IRB (now Advarra).

### Summary of Updates

This version used the name naltrexone and acetaminophen combination rather than naltrexone/acetaminophen.

